# Modeling-based UV-C decontamination of N95 masks optimized to avoid undertreatment

**DOI:** 10.1101/2020.10.30.20223354

**Authors:** Adam P. Sears, Jacques Ohayon, Anton D. Shutov, Roderic I. Pettigrew

## Abstract

As the Coronavirus 2019 pandemic creates worldwide shortages of personal protective equipment, hospitals have increasingly turned to sterilization and re-use protocols, often without significant data supporting the specific methodologies. When using UV-C irradiation, previously shown to be effective for decontaminating hard surfaces, modeling shows the importance of accounting for the porosity and non-uniform curvature of the N95 masks in decontamination procedures. Data shows a standard incident dose of 1 J/cm^2^ delivered to both front and back surfaces is more than 500x higher than the known kill dose. However, modeling indicates this would undertreat 40% of the mask material due to the curvature, path-length attenuation and scatter. Multidirectional UV-C irradiation employing dose calibrated exposures can adjust for this loss and best decontaminate masks. Such protocols can be rapidly implemented in thousands of hospitals across the world equipped with UV-C irradiation lamps without the need for additional capital equipment purchases.

## Introduction

The Coronavirus disease-2019 (COVID-19) has sickened over 6 million people around the world, with several hundred thousand hospitalizations. The rapid influx of patients to hospitals and intensive care units has resulted in a high demand for personal protective equipment (PPE) with constant supply-chain pressure leading to shortages and rationing. Fitted N95 masks which block aerosolized virus-containing droplets are of particular importance in protecting clinicians and front-line workers from being infected with and spreading this dangerous pathogen. While traditionally considered single-use items, some research has looked at and tested the possibility of sanitizing masks for re-use [1–5]. This is not yet a common practice as techniques are evolving and the availability of necessary equipment is likely site specific.

Protocols for hospital sterilization frequently use ultraviolet C (UV-C) germicidal irradiation to sanitize patient rooms and surgical suites. UV-C irradiation has gained favor since it is rapid and effective for sterilizing hard surfaces, making it an ideal choice for repeated sanitization of rooms [6]. Light at wavelengths within the UV-C band inactivate viruses through DNA and RNA lysis. However, N95 masks and surgical masks are engineered with complex geometries and with layers of porous material. This creates layers within the thickness of the mask material where germs or viral particles might penetrate and reside. If the UV-C light does not sufficiently penetrate the mask material, the viral particles and infectious agents that can become embedded within the layers of N95 masks may be untreated by traditional UV-C sterilization protocols designed for treating hard surfaces.

Because UV-C light is highly absorbed by most material, this attenuation–an exponential function of material thickness and type–and some degree of light scatter must be considered in effective treatment planning for masks sterilization using UV-C light. Adaptation of existing UV-C systems to this purpose [7, 8], however, allows mask sterilization to be done at the point of care, employing equipment already within many hospitals around the world. Here we report simulations of treatment with a focus on the sanitization of the entire mask, including interior layers. Our findings highlight several issues with simple illumination configurations [1] while describing an efficient protocol for operation of UV-C treatment of masks.

## Materials and methods

### Setup

We used mathematical and computational modeling to predict the attenuation of UV-C light rays in a decontamination apparatus where masks are suspended along parallel lateral supports (Fig 1) with UV-C lamps placed on either side. We considered a treatment array that is 200 cm wide x 100 cm high and targeted a minimum UV-C dose of 20 mJ/cm^2^ [9, 10], up to 4x the decimation dose for viruses such as SARS-CoV-2, across all masks within the array, shown in Fig 1a.

**Fig 1.**
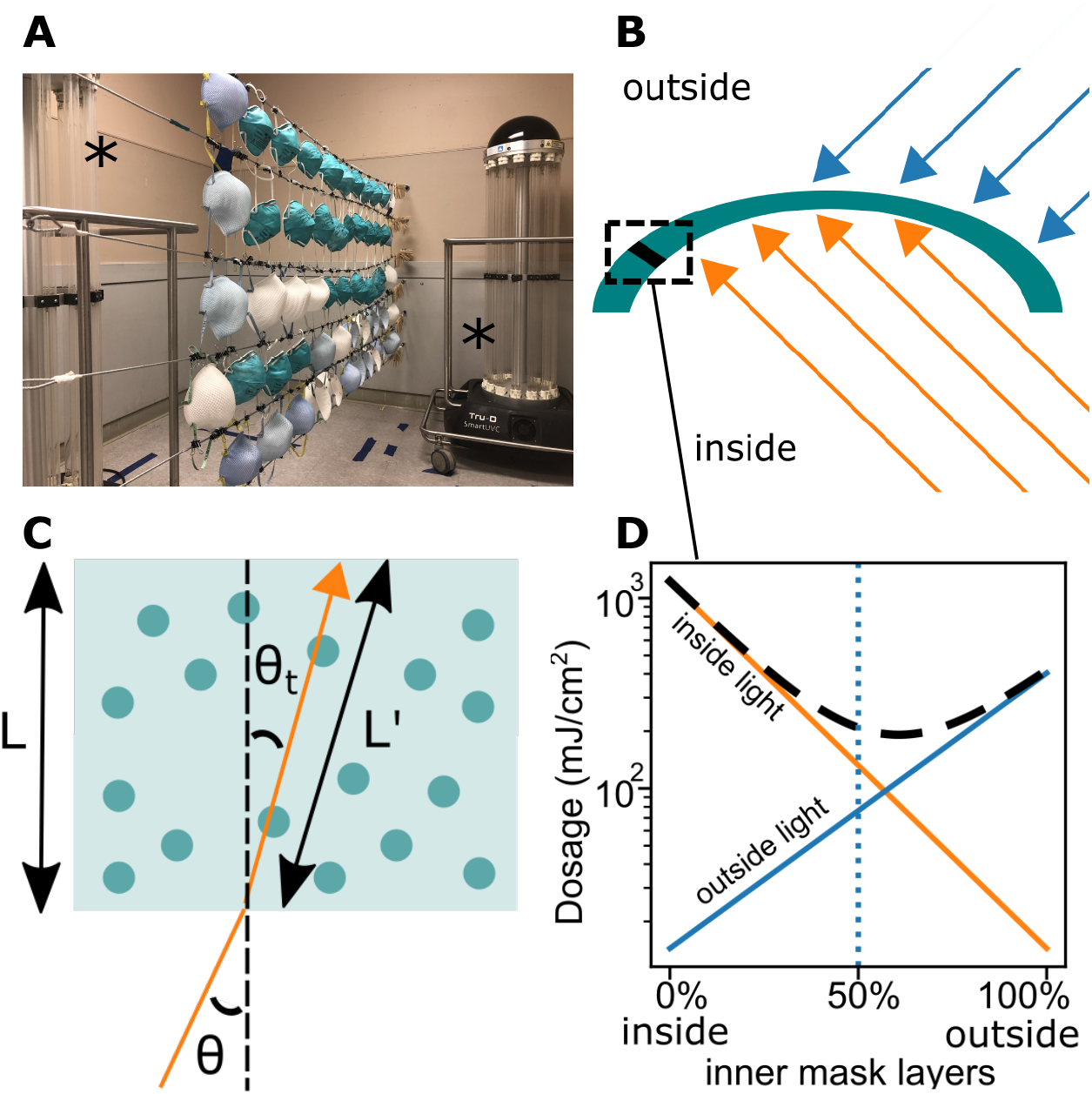
a) The apparatus setup, showing a treatment array and two lamps marked by (*) b) a curved mask exposed to one UV-C lamp on each side. Due to the mask curvature, some areas are not directly exposed to the light rays of the two lamps. c) UV-C light ray (orange arrow) is refracted and penetrates the mask panel fabric. Attenuation of light at a given depth increases via scattering or with larger incidence angle *θ* d) UV-C dosage is attenuated as light enters the inner layers of the N95 mask fabric, due to the effects of mask curvature on light attenuation

We used Tru-D UV-C robots (Tru-D SmartUVC, Memphis, TN), designed for room irradiation and sterilization, to illuminate the masks, and calibrated the total intensity of each lamp using a Gigahertz-Optik X11-1-UV3718 radiometer (Turkenfeld, Germany). Lamps typically produced 2.7 J/cm^2^ over 5 minutes of exposure at a distance of 55 cm and an elevation of 90 cm, half the height of the lamp. Treatment dose is additive and any number of lamps may be used, although we used one lamp on each side of the treatment array while performing protocols with either single or multiple stations.

Masks may be more difficult to treat depending on their curvature and layered composition. We chose to perform our simulations using the 3M model 1860 because we found it more challenging to decontaminate than other N95 masks such as the Moldex model 2200. The Moldex N95 mask has comparable curvature and thickness, but is significantly more transparent to UV-C light than the 3M version, which improves exposure of the mask interior. We selected Halyard tie-back masks as a representative flat folded surgical mask.

### Modeling

We modeled the dose received by each part of a medical mask by considering the emission of ultraviolet light from a UV-C lamp, the path it took to reach its mask target, and the angle of incidence *θ* at the mask surface. Energy emitted from the UV-C lamps is assumed approximately uniform over all angles from each section of its mercury vapor bulbs. Lamps are composed of 14 tubes, each of which was discretized into 20 light sources. Therefore each mask segment receives 14×20 light rays of distinct orientations per lamp position. Masks positioned a distance *R* away from the light source receive an ultraviolet dose reduced by a factor of 1*/R*^2^ at their surface.

For a given segment of a mask on the treatment rack, the angle of incidence of each light ray entering the fabric is cos(*θ*) = *L* · *M* where *L* is the unit light-vector from the tube section to the mask segment, and *M* is the segment unit normal vector. The density of light on the fabric decreases with distance *R*, but the attenuation of light as it travels within the porous mask fabric is much more significant, and this loss is enhanced with larger *θ* as the path length increases. We assumed the index of refraction of the mask n *≈* 1.5 [11] and included surface reflection and refraction of transmitted rays described by Snell’s law [12], which describes *θ*_*t*_, the transmitted angle using the relation sin(*θ*) = *n* sin(*θ*_*t*_). Light is attenuated exponentially as it passes through the multiple layers of the fabric, and the energy density at depth *x* inside the mask segment is the sum of components [13]:

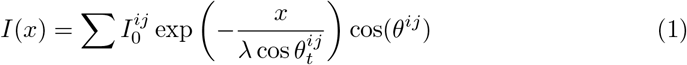

where for the light ray *i* originating from lamp tube 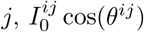 are the energy density just inside the mask and normal to its surface, *θ*^*ij*^ are the angle of incidence, 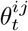 are the transmitted angles, and *λ* is a material characteristic decay length. We determined *λ*, assumed to be constant in the fabric, through experiments at *θ* = 0 with and without the mask fabric.

The internal intensity is the sum of all rays, including exposures from multiple lamps or lamp positions. Masks placed at some locations have receive more or less treatment due to the relative strengths and decay (attenuation and scatter) of these rays. Data deposition: code used for simulation will be available from the GitHub repository (https://github.com/PettigrewLab/N95Simulation.git) upon article publication.

### Optimization

We considered mask treatment at each point of the array where a mask could be placed. This involved simulated exposure along a 3M N95 mask’s largest cross-sectional profile, assuming quadratic curvature. In Fig 1 these values are reduced to a single pixel by taking the minimum along this cross-section, while the full profile is presented in Fig 2. As a baseline, we consider a simple protocol for these figures, where the masks are exposed until a sensor at the bottom edge of the array accumulates a 300 mJ/cm^2^ dose [1]. However because a single pair of lamp positions required impractical exposure times to produce adequate treatment, we also designed a protocol which used two pairs of locations. To optimize this, we maximized the minimum dose deposited in each mask across the array, through varying both the longitudinal and lateral distances of the lamp locations. This achieves the fastest treatment and the highest system throughput.

**Fig 2.**
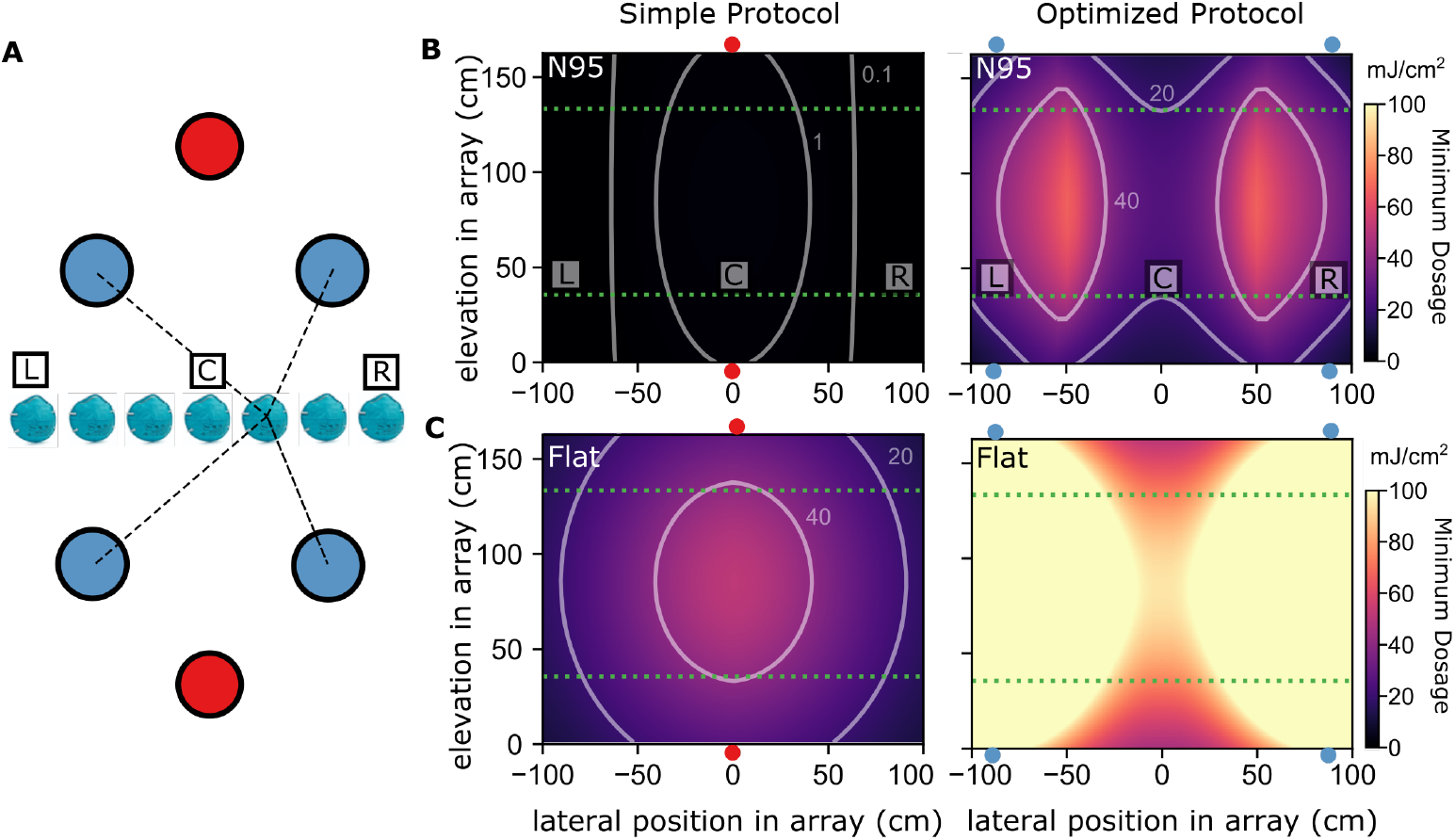
The numerical results from the optimized protocol are compared to those obtained by simulating the protocol used by Lowe et al. [1]. a) Diagram of a set of medical masks in front of UV-C lamps placed symmetrically on each side of the array of masks (viewed from top / above). Two decontamination protocols using different lamp locations are presented: (red) a simple protocol [1], and (blue) an optimized protocol with 4 locations. L, C and R are respectively the left-most, center and right-most masks at the bottom of the mask array. b) Minimum dose received for decontamination protocols by any portion of a mask, when placed at a given lateral position and elevation (left) A simple protocol fails to treat the array fully (right) The optimized protocol with 2 pairs of lateral locations adequately treats all masks within the array (the green dashed lines) after time of 15 minutes per lamp location. c) Minimum dose received by each mask for a set of flat-folded surgical masks. (left) The simple protocol treats 87% of the 200 cm-wide x 100 cm-high treatment array. (right) The optimized protocol has no difficulty treating flat masks in all positions of the array.

## Results

In Fig 1 we compare a simple protocol using a single pair of lamps against an optimized protocol that uses illuminations from two pairs of lamps. Simulations showed marked variation in the transmitted UV-C levels across masks, highly dependent upon their curvature and the position of the mask relative to the light source. The contour plot shows that the simpler arrangement does not treat N95 masks effectively, and would require infeasible exposure times to treat masks along the entire array. Portions of masks at every position received less than even a single-decimation dosage of 2 mJ/cm^2^, with some surfaces not exposed directly due to mask curvature. In an optimized protocol, however, adequate decontamination can be achieved by using our lamps for 15 minutes at each of the two optimized paired positions along the array (Fig 2). In contrast, flat folded masks can be effectively treated even with the simple protocol, with masks on over 87% of the array receiving the desired dose throughout their entirety.

The way in which the simple protocol fails for curved masks is illustrated in Fig 3, in which simulated treatment dosage within individual masks is displayed. When the masks are exposed to light from only a single direction on each side, there is an order of magnitude more variance UV-C dosage throughout the mask volume. Masks positioned at extreme angles receive very unequal exposure and some parts may only be illuminated on one side. Optimized exposure from two directions leads to a much more uniform dose. At lateral lamp positions of *±*91 cm, placed a longitudinal distance 52 cm from the array, a 2×2 lamp protocol is optimal and requires about 15 minutes exposure per lamp, or about 30 minutes total with two robots.

**Fig 3.**
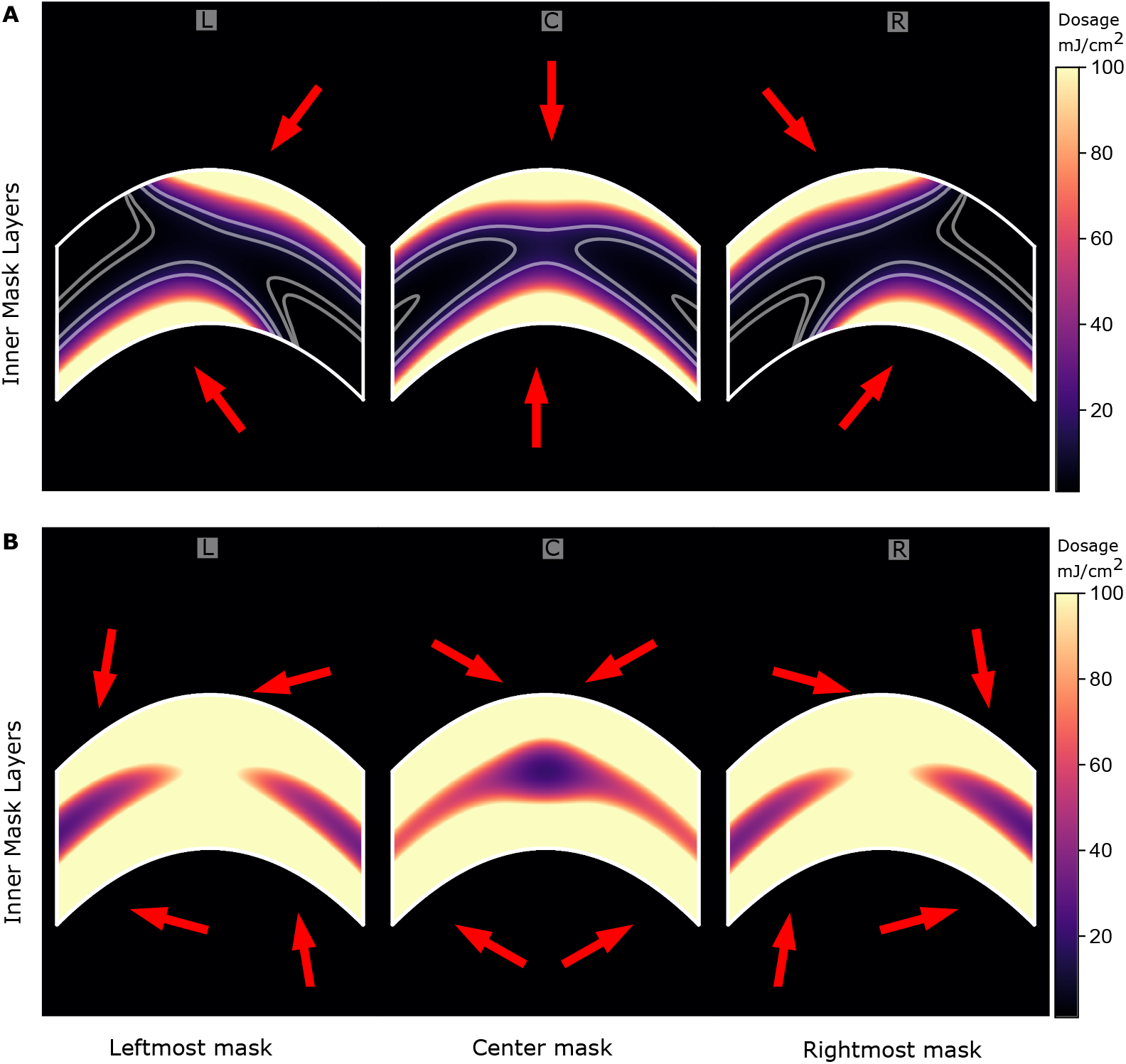
Penetration light intensity in the inner layers of the N95 mask fabric, for the simple and optimized protocols. The internal dose distributions are shown for the left-most (L), center (C) and right-most (R) masks at the bottom of the treatment array. The results from the optimized protocol are compared to those obtained when using a single lamp centered on both sides of the array. Red arrows indicate the general orientation of the light from each light source. a) The simple protocol (2 red lamps), does not effectively treat the inner layers of the mask fabric. The interior and some external surfaces of masks located at the ends of the planar array are not properly decontaminated. b) The optimized 2×2 protocol (4 blue lamp positions), sterilizes the masks with a more uniform dose distributions for masks throughout the array.

## Discussion

For masks that cannot be flattened, the attenuation becomes more important with larger angles of incidence, and largely determines the necessary irradiation time, which must be set so that the delivered dose throughout every mask within the treatment array reaches a minimum level. Light that reaches the mask surface from the UV-C source is further absorbed by the mask material at depth (Fig 3). Incident energy which reached the reverse side of these masks was decreased by a factor of 1000x for the 3M N95 mask. The light transmission drop for a Moldex mask was substantially lower with a reduction factor of approximately 100x, while flat-folded surgical masks had an opacity between these two values. Mask curvature cannot be neglected and leads to undertreatment of inner layers.

The minimum dose within the mask layers, across the mask surface, and along the mask array, determines the effectiveness of treatment. Despite the symmetry of illuminating both the front and back, one side or surface of a curved mask which is illuminated obliquely can limit treatment effectiveness (Fig 2). However, exposure from multiple angles mitigates imbalance and is ultimately more efficient than longer exposure time to reach a target dose.

Another common heuristic, targeting a surface intensity dosage of 1 J/cm^2^ (on both sides of the N95 mask [7]) undertreats a 20 mJ/cm^2^ target for nearly 40% of the material in N95 masks. To efficiently achieve a dosage of 20 mJ/cm^2^, 4-10x the inactivation dose for viruses [9, 10] similar to the novel coronavirus SARS-CoV-2, it is critical to know the minimum dose being delivered to each portion of the mask which may be contaminated. We found that, throughout the full mask material, the delivered dose distribution for any single mask showed a large variance, differing by more than 100x. This is primarily due to the curvature and difference in incident light path lengths for locations across the mask when illuminated from a single source on both sides (Fig 3). Increasing the number of exposure angles is an efficient way of lowering the total treatment time, and using the optimized configuration with two positions on each sides of the mask reduces dosage variance to ≈2.5x (Fig 1). There are diminishing returns from using 3 paired locations, with only slightly greater uniformity and efficiency at this array size.

Re-using masks helps to mitigate potential dangerous PPE shortages that would leave clinicians and communities vulnerable to a viral pandemic. UV-C sterilization using multiple angle exposures to ensure mask safety is a feasible option for the many hospitals across the world that already have mobile UV-C systems. Hundreds of these masks can be processed in a day with a single suspension system. However, care should be taken to ensure sanitization of the mask interior layers, which is highly sensitive to mask curvature and has been neglected in previous work. Implementation of properly designed protocols could prevent the need for additional capital investment from hospitals already incurring reduced revenue and increased expenses.

## Data Availability

Code will be released upon journal publication

https://github.com/PettigrewLab/N95Simulation

## Acknowledgments

The authors thank Kristen Maitland for discussions on utilization of UV-C irradiation for decontamination and Erika Nolte for assistance with manuscript preparation. We are also grateful for the efforts and support of Daniel Felan and Mohamed Fawaz.

